# Association of Bleeding Risk With Recurrent Cardiovascular Events in Asian Patients with Acute Coronary Syndrome: A Nationwide Population-Based Cohort Study

**DOI:** 10.1101/2024.10.10.24315273

**Authors:** Chih-Wei Chen, Yi-Cheng Lin, Donna Shu-Han Lin, Chia-Li Chang, Chun-Yao Huang, Jaw-Wen Chen, Shing-Jong Lin, Yu-Hsuan Shao, Chien-Yi Hsu

**Affiliations:** Division of Cardiology and Cardiovascular Research Center, Department of Internal Medicine, Taipei Medical University Hospital, Taipei, Taiwan; Department of Medical Research, Taipei Medical University Hospital, Taipei, Taiwan; Department of Pharmacy, Taipei Medical University Hospital, Taipei, Taiwan; Clinical Big Data Research Center, Taipei Medical University Hospital, Taipei, Taiwan; Division of Cardiology, Department of Internal Medicine, School of Medicine, College of Medicine, Taipei Medical University, Taipei, Taiwan; Taipei Heart Institute, Taipei Medical University, Taipei, Taiwan; School of Pharmacy, College of Pharmacy, Taipei Medical University, Taipei, Taiwan; Graduate Institute of Biomedical Informatics, Taipei Medical University, Taipei, Taiwan; Health Data Analytics and Statistics Center, Office of Data Science, Taipei Medical University, Taipei, Taiwan; Graduate Institute of Clinical Medicine, Taipei Medical University, Taipei, Taiwan; Division of Cardiology, Department of Internal Medicine, Shin Kong Wu Ho-Su Memorial Hospital, Taipei, Taiwan

## Abstract

**Background:** Acute coronary syndrome (ACS) carries significant risks of recurrent cardiovascular (CV) events and bleeding complications. In particular, Asian patients have higher rates of bleeding complications due to genetic and physiological factors. Bleeding complications are associated with an increased risk of subsequent thrombotic events, and the impact of such complications on long-term outcomes must therefore be investigated. This study compared long-term outcomes and clinical characteristics between ACS patients who experienced a single ACS event and those who experienced multiple CV events.

**Methods:** Utilizing data from Taiwan’s National Health Insurance Research Database, this retrospective cohort study categorized patients into single-event and multiple-event groups based on the occurrence of major adverse CV events within 2 years after the index ACS event. In this cohort study, 28,535 patients were included. After matching by age, sex, and the interval between the first and second CV events at a 1:2 ratio, 8,720 patients were included in the multiple-event group and 17,368 in the single-event group.

**Results:** The multiple-event group had higher rates of comorbidities, including hypertension, prior coronary artery disease, heart failure, stroke, and chronic kidney disease. Over a 5-year period, the multiple-event group exhibited higher all-cause mortality (34.1% vs. 24.6%, p < 0.0001) and CV mortality (11.4% vs. 6.2%, p < 0.0001) than the single-event group. The rates of major bleeding events (7.8% vs. 1.6%, p < 0.0001) and minor bleeding events (34.4% vs. 7.2%, p < 0.0001) were also higher in the multiple-event group than in the single-event group. Compared with the single-event group, which showed a significant reduction in major bleeding events 1 month after the index ACS event, the multiple-event group continued to have a higher rate of major bleeding events within 3 months following the index ACS event. In the multiple-event group, patients who experienced a major bleeding event had an earlier onset of subsequent CV events than patients who did not experience a major bleeding event. Specifically, every 1-day earlier occurrence of major bleeding was associated with a 1.0044-day earlier occurrence of a subsequent CV event.

**Conclusion:** ACS patients with multiple CV events have higher rates of all-cause mortality, CV mortality, and major bleeding than ACS patients with a single CV event. However, major bleeding may be associated with the risk of subsequent CV events, highlighting the importance of implementing a tailored antiplatelet strategy in Asian populations.

## Introduction

Cardiovascular (CV) disease is one of the main causes of mortality in developed countries, including countries in Asia.^1,2^ In recent years, with progress in acute coronary syndrome (ACS) treatment, including advanced reperfusion techniques and innovative antithrombotic treatments, the approach to the management of ACS has undergone substantial changes, leading to considerable improvements in patient outcomes.^3,4^ Despite advances in medical treatment, ACS still carries a very high risk of recurrent ischemic events.^5^ Research indicates that the recurrence rate of acute myocardial infarction (AMI) within 1 year after the initial myocardial infarction event approaches 10%. Additionally, the incidence of CV events remains high in the subsequent years.^6,7^ Recurrent CV events result in cardiac function impairments, which may increase the risk of heart failure, adversely affect the patient’s quality of life, and result in death.^8^

There is a relative paucity of research on bleeding risks in patients with multiple cardiovascular events. While some studies indicate that high ischemic risk, as defined by the European Society of Cardiology/European Association for Cardio-Thoracic Surgery (ESC/EACTS) guidelines, does not necessarily correlate with increased clinically relevant bleeding risks.^9^ However, this issue warrants further investigation, particularly in the era of widespread use of new P2Y12 inhibitors (e.g., ticagrelor or prasugrel). Concerns are rising regarding elevated bleeding risks among Asian patients with multiple cardiovascular events, making it crucial to examine this issue more thoroughly in the context of contemporary treatment practices.^10^

Therefore, this study compared long-term outcomes and clinical characteristics between ACS patients who did not experience subsequent CV events (the single-event group) and ACS patients who experienced additional CV events (the multiple-event group). This comparison was conducted to elucidate the impact of multiple CV incidents on patient prognosis and ACS management strategies.

## Methods

### Data and patients

This retrospective population-based cohort study analyzed data from Taiwan’s National Health Insurance Research Database (NHIRD), which is a comprehensive, nationwide, claim-based repository encompassing the reimbursement claims of insurants covered by the National Health Insurance (NHI) program. The database contains various types of health-care data, including patient enrollment details, diagnostic codes from the International Classification of Diseases, Ninth and Tenth Revisions (ICD-9 and ICD-10, respectively), codes for medical procedures, and medication records from hospital admissions and outpatient visits. The present study received ethical approval from Taipei Medical University’s Joint Institutional Review Board (TMU-JIRB No.202305022). Because data from the NHIRD are anonymized, the requirement for informed consent was waived by the Taipei Medical University’s Joint Institutional Review Board.

The study population included patients aged 20 years and older who had survived, had a discharge record of ACS, and was treated with anticoagulant, aspirin, or a P2Y12 inhibitor or an GPIIb/IIIa antagonist from 2013 to 2015. Participants were categorized into two groups: the single-event group, consisting of patients who experienced only one ACS episode within 2 years, and the multiple-event group, including patients who had additional CV events (including ACS requiring PCI or CABG or ischemic stroke) within 2 years after the initial ACS event. The detailed inclusion criteria are listed in the **supplementary data file**.

### Outcome

The primary outcome of this study was all-cause mortality (including CV mortality and other causes of death), which was verified against data from the National Death Registry. The secondary outcomes were major and minor bleeding events. The detailed definitions of the outcomes are provided in the **supplementary data file**.

### Statistical analysis

The patients in the single-event group and the multiple-event group were matched by age, sex, and the interval between the first and second events (index date) at a 1:2 ratio. The index date for the multiple-event group was the date of the second ACS event, and the index date for the single-event group was the date of the second event in the matched patient from the multiple-event group. Clinical characteristics were compared between the two groups by using the chi-square test for categorical variables and the *t* test for continuous variables. The follow-up period was defined as the time from the index date until either the occurrence of death or the end of the study period, whichever occurred first. A conditional survival analysis was used to analyze clinical characteristics in relation to primary and secondary outcomes. The results are presented as hazard ratios (HRs) with 95% confidence intervals. In addition, event rates and adjusted HRs for all-cause mortality, CV mortality, and bleeding events were assessed in relation to the number of risk factors. Data analyses were performed using SAS statistical software, version 9.4 (SAS Institute Inc., Cary, North Carolina, USA). A p value of <0.05 indicated statistical significance.

### Results

In this cohort study, 28,535 patients were included. After the exclusion of 1) patients aged <20 years, 2) individuals with no data on sex, and 3) individuals who did not survive during hospitalization, 26,833 patients remained eligible for analysis. Of these, 8,721 patients (32.5%) were classified into the multiple-event group and 18,112 patients (67.5%) into the single-event group. Following matching by age, sex, and the interval between the first and second CV events at a 1:2 ratio, 8,720 patients were included in the multiple-event group (33.49%) and 17,368 patients in the single-event group (66.56%) (**Figure 1**).

**Figure 1.**
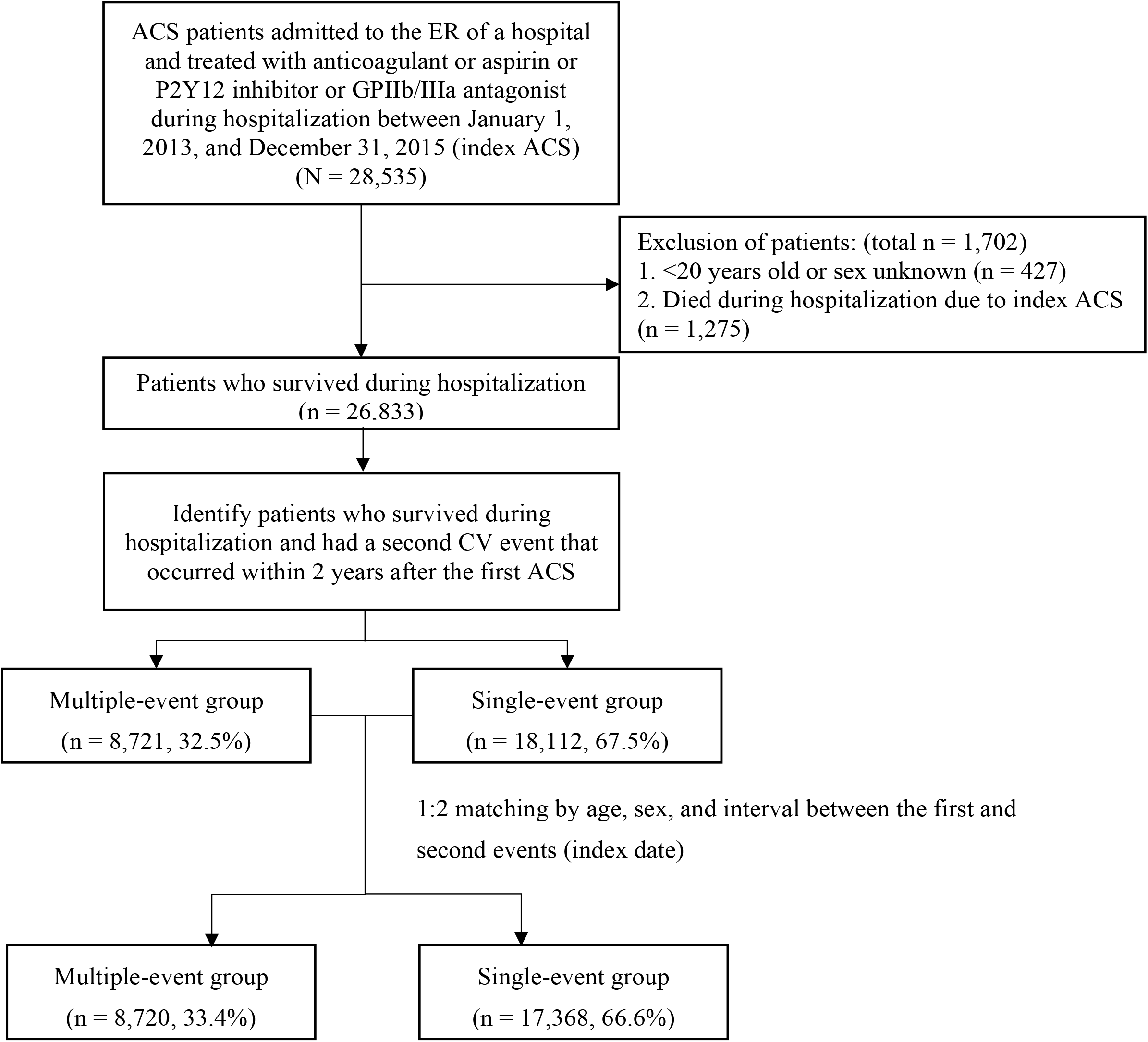
Flowchart of the selection of patients with ACS The index date was defined as the date of the second CV event for the multiple-event group and the corresponding date for the single-event group.

The baseline characteristics for both groups are presented in **Table 1**. Following matching by sex and age, the multiple-event group had a higher rate of comorbidities, including hypertension, prior coronary artery disease (CAD), heart failure, stroke, atrial fibrillation, peripheral vascular disease, and chronic kidney disease (CKD). Prior to the index ACS event, a higher proportion of patients in the multiple-event group had received treatment with aspirin, P2Y12 inhibitors, and other CV medications such as beta-blockers, ACE inhibitors/angiotensin receptor blockers (ACEI/ARB), and statins.

**Table 1.** Baseline characteristics of ACS patients ^a^ (N = 26,088)

| Characteristics | Single-event group |  | Multiple-event group |  | SMD<sup>b</sup> | p value<sup>c</sup> |
| --- | --- | --- | --- | --- | --- | --- |
|  | (n = 17,368) |  | (n = 8,720) |  |  |  |
|  | n | (%) | n | (%) |  |  |
| Age at index, year |  |  |  |  |  |  |
| Mean ± SD | 64.7 ± 13.4 |  | 64.7 ± 13.5 |  | 0.000 | - |
| Median (IQR) | 64.4 (55.2 - 75.3) |  | 64.4 (54.9 - 75.3) |  |  |  |
| Male | 13,406 | (77.2) | 6,721 | (77.1) | 0.003 | - |
| Index ACS |  |  |  |  |  |  |
| STEMI | 14,081 | (81.1) | 7,083 | (81.2) | 0.004 | 0.3916 |
| NSTEMI | 1,340 | (7.7) | 700 | (8.0) | 0.012 |  |
| Unstable angina | 1,947 | (11.2) | 937 | (10.7) | 0.015 |  |
| Comorbidity |  |  |  |  |  |  |
| Diabetes | 5,026 | (28.9) | 3,387 | (38.8) | 0.210 | <0.0001 |
| Hyperlipidemia | 4,553 | (26.2) | 2,668 | (30.6) | 0.097 | <0.0001 |
| Gout | 1,498 | (8.6) | 827 | (9.5) | 0.030 | 0.0216 |
| Hypertension | 8,654 | (49.8) | 5,136 | (58.9) | 0.183 | <0.0001 |
| CAD | 3,228 | (18.6) | 2,580 | (29.6) | 0.259 | <0.0001 |
| Heart failure | 1086 | (6.3) | 898 | (10.3) | 0.147 | <0.0001 |
| Stroke | 819 | (4.7) | 991 | (11.4) | 0.246 | <0.0001 |
| Atrial fibrillation | 457 | (2.6) | 308 | (3.5) | 0.052 | <0.0001 |
| Peripheral artery disease | 266 | (1.5) | 246 | (2.8) | 0.088 | <0.0001 |
| Ventricle arrhythmia | 27 | (0.2) | 21 | (0.2) | 0.019 | 0.1291 |
| Chronic lung disease | 1,884 | (10.8) | 972 | (11.1) | 0.010 | 0.4652 |
| CKD | 1,777 | (10.2) | 1,511 | (17.3) | 0.207 | <0.0001 |
| Cancer | 872 | (5.0) | 400 | (4.6) | 0.020 | 0.1250 |
| Medications |  |  |  |  |  |  |
| PPI | 496 | (2.9) | 379 | (4.3) | 0.080 | <0.0001 |
| Insulin | 795 | (4.6) | 882 | (10.1) | 0.213 | <0.0001 |
| Oral anticoagulant | 322 | (1.9) | 174 | (2.0) | 0.010 | 0.4301 |
| P2Y12 inhibitor | 405 | (2.3) | 846 | (9.7) | 0.314 | <0.0001 |
| Aspirin | 3,048 | (17.5) | 2,386 | (27.4) | 0.237 | <0.0001 |
|  | (n = 17,368) |  | (n = 8,720) |  |  |  |
|  | n | (%) | n | (%) |  |  |
| Beta-blocker | 3,481 | (20.0) | 2,403 | (27.6) | 0.177 | <0.0001 |
| CCB | 3,996 | (23.0) | 2,548 | (29.2) | 0.142 | <0.0001 |
| ACEI or ARB | 5,457 | (31.4) | 3,536 | (40.6) | 0.191 | <0.0001 |
| Statins | 3,186 | (18.3) | 2,198 | (25.2) | 0.167 | <0.0001 |
| NSAID | 1,896 | (10.9) | 886 | (10.2) | 0.025 | 0.0620 |
| Urate lowering drugs | 1,421 | (8.2) | 897 | (10.3) | 0.073 | <0.0001 |
| PCI or CABG at index ACS event | 12,490 | (71.9) | 6,525 | (74.8) | 0.066 | <0.0001 |

<sup>a</sup> Match by age, sex, and the interval between the first and second events (index date). The index date was the date of the second CV event for the multiple-event group and the corresponding date for the single-event group. <sup>b</sup> Standardized mean difference (SMD) indicates the difference in means or proportions divided by the standard error; imbalance is defined as an absolute value greater than 0.1. <sup>c</sup> Chi-square test or independent *t* test.

In terms of the outcomes displayed in **Table 2** over a 5-year follow-up period, the multiple-event group exhibited higher incidence rates of all-cause mortality (34.1% vs. 24.6%, p < 0.0001), CV death (11.4% vs. 6.2%, p < 0.0001), and non-CV death (22.7% vs. 18.3%, p < 0.0001) than the single-event group. **Figure 2** depicts the Kaplan–Meier curves for all-cause mortality and CV death. The multiple-event group exhibited higher rates of major bleeding events (7.8% vs. 1.6%, p < 0.0001) and minor bleeding events (34.4% vs. 7.2%, p < 0.0001) than the single-event group. **Figure 3** illustrates the Kaplan–Meier curves for major bleeding and minor bleeding events.

**Table 2.** Comparative mortality and bleeding events in single-vs. multiple-cardiovascular-event groups.

|  | Single-event group |  | Multiple-event group |  | SMD<sup>a</sup> | p value<sup>b</sup> |
| --- | --- | --- | --- | --- | --- | --- |
|  | (n = 17,368) |  | (n = 8,720) |  |  |  |
| Event | n | (%) | n | (%) |  |  |
| All-cause death | 4,267 | (24.6) | 2,975 | (34.1) | 0.211 | <0.0001 |
| CV death | 1,084 | (6.2) | 998 | (11.4) | 0.184 | <0.0001 |
| Other-cause death | 3,183 | (18.3) | 1,977 | (22.7) | 0.108 | <0.0001 |
| Bleeding |  |  |  |  |  |  |
| Major | 281 | (1.6) | 684 | (7.8) | 0.296 | <0.0001 |
| Minor | 1,256 | (7.2) | 2,998 | (34.4) | 0.710 | <0.0001 |

<sup>a</sup> Standardized mean difference (SMD) indicates the difference in means or proportions divided by the standard error; imbalance is defined as an absolute value greater than 0.1. <sup>b</sup> Chi-square test or independent *t* test.

**Figure 2.**
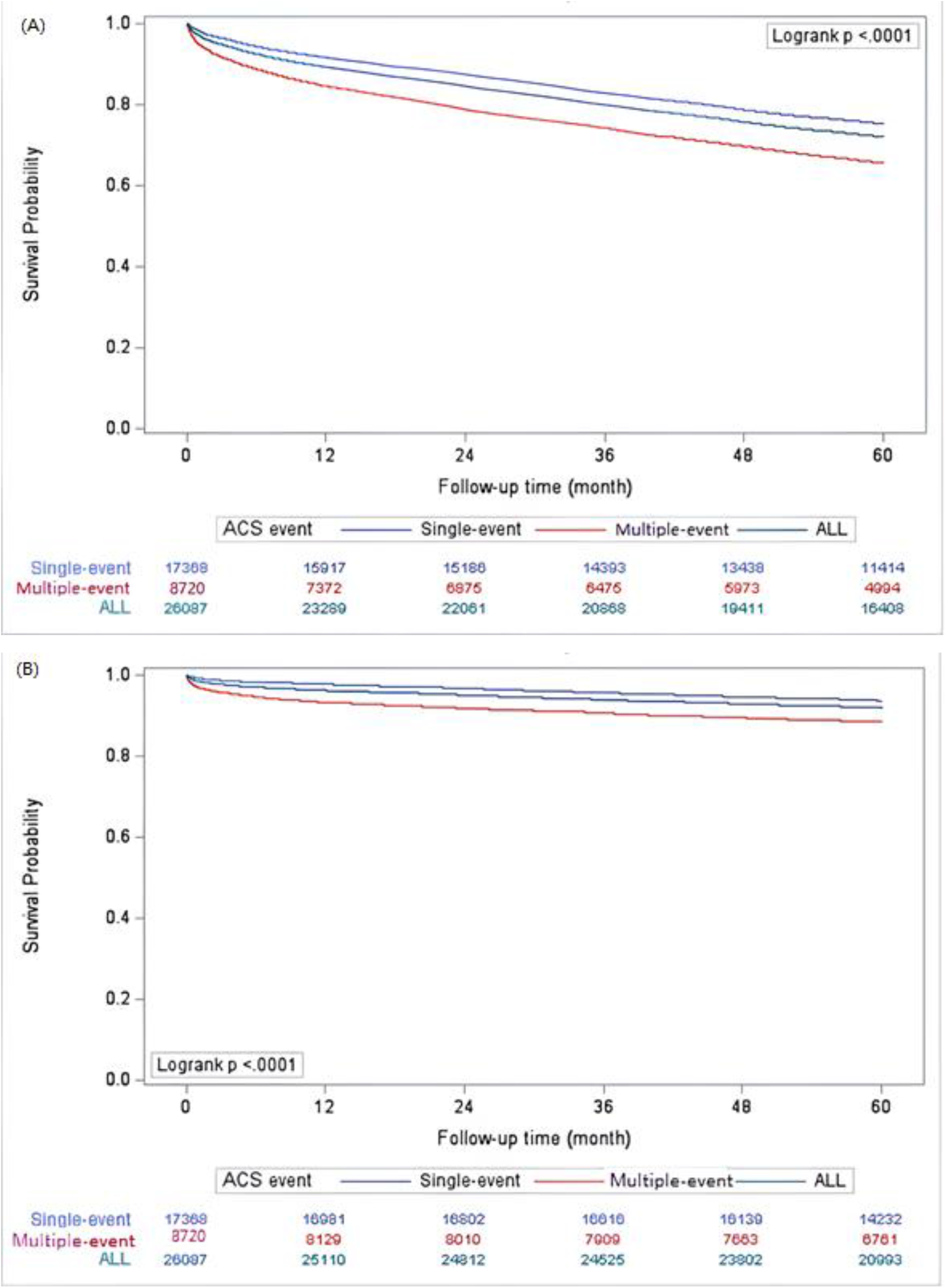
Survival probability of ACS patients after the index date in terms of (A) all-cause mortality and (B) CV death

**Figure 3.**
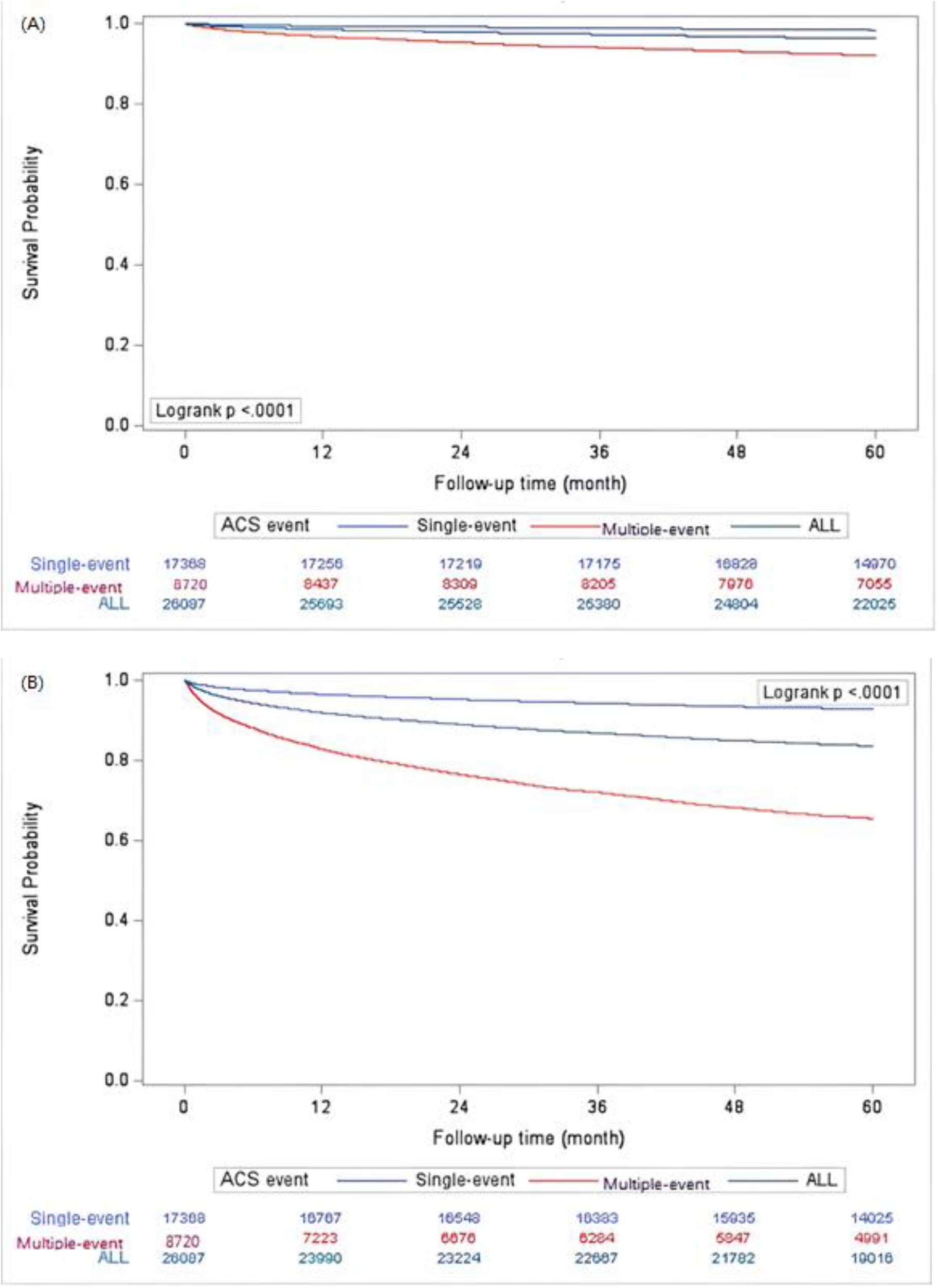
Survival probability of ACS patients after the index date in terms of (A) major bleeding and (B) minor bleeding.

**Figure 4.**
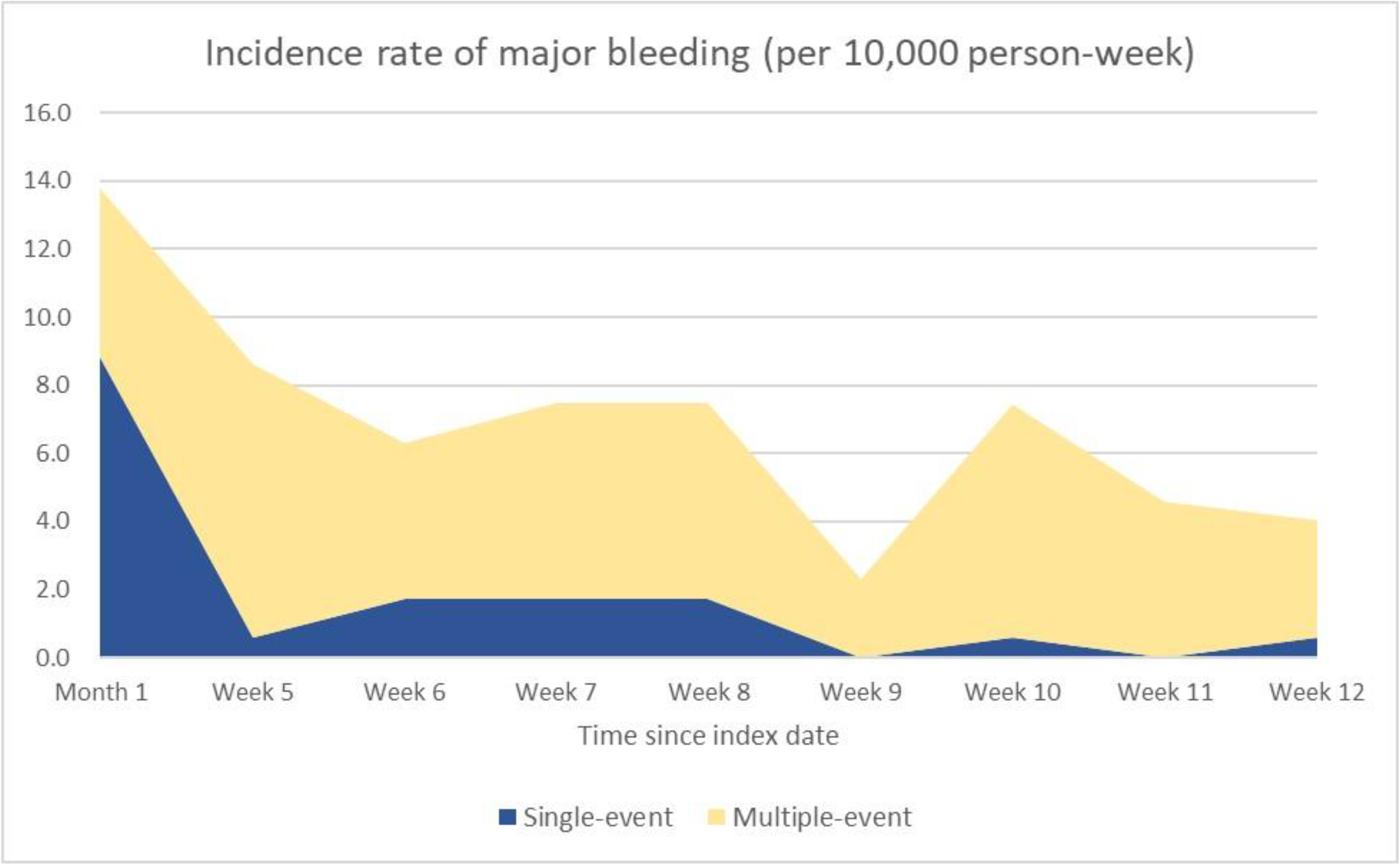
Incidence rate of major bleeding within 3 months after the index date

Upon further analysis for the multiple-event group, among patients who experienced a major bleeding event, every 1-day earlier occurrence of major bleeding was associated with a 1.0044-day earlier onset of a subsequent CV event (Table 3).

**Table 3.**
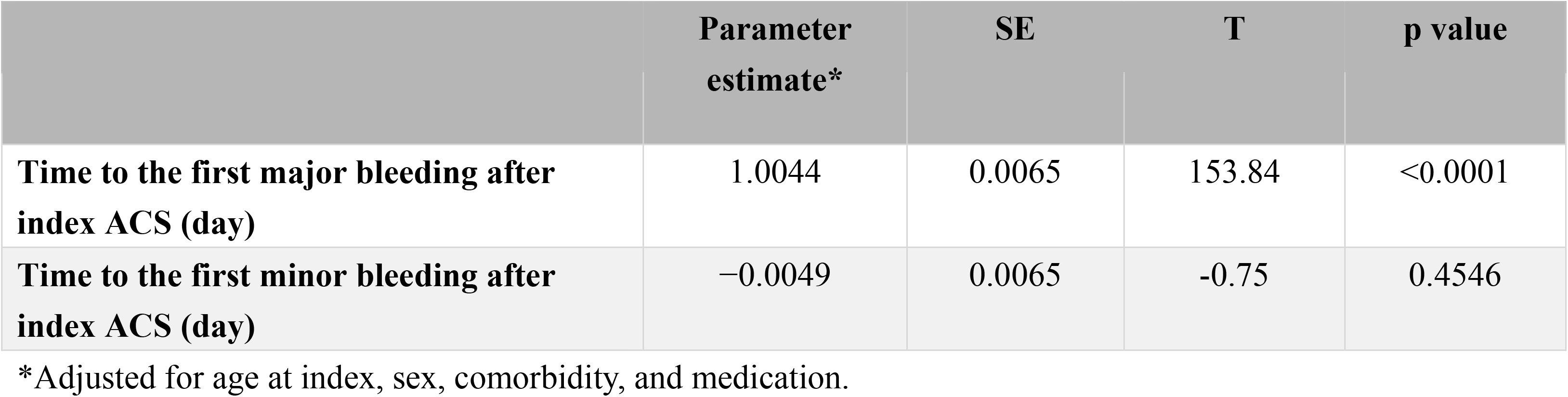
Relationship between time to a subsequent CV event (day) and time to bleeding after index ACS in the multiple-event group (n = 8,720)

## Discussion

This retrospective cohort study demonstrated that after matching by age and sex, ACS patients with multiple CV events exhibited a higher all-cause mortality rate than ACS patients with a single ACS event, including both CV and non-CV deaths. Additionally, ACS patients who experienced multiple CV events exhibited higher rates of major and minor bleeding during follow-up.

Numerous studies have indicated that patients with recurrent myocardial infarction exhibit poorer outcomes than patients with new-onset myocardial infarction.^11,12^ This finding is consistent with ours. Diabetes and CKD have been identified as significant predictors of the recurrence of myocardial infarction.^13-15^ Similarly, our study also revealed that the multiple-event group had a higher prevalence of comorbidities such as diabetes and CKD. In our study, nearly one-third of ACS patients (32.5%) experienced an additional CV event within 2 years after the index ACS event, including PCI, stroke, or another ACS episode. A meta-analysis of four randomized controlled trials found a CV event rate of approximately 9.2% in the first year following the initial ACS event, with rates continuing to escalate in the second and third years of follow-up.^16^ Our data on baseline characteristics revealed that patients in the multiple-event group had higher rates of comorbidities and were more likely to have been receiving CV medications, such as anticoagulants, aspirin, P2Y12 inhibitors, and statins, prior to the index ACS event. Compared with the single-event group, the multiple-event group had a higher incidence of all-cause death and bleeding events, including major and minor bleeding. These results are particularly interesting in a cohort predominantly composed of Asian people, and they echo previous findings, such as those of the TALOS-AMI and AFIRE trials. These trials have demonstrated that the excessive use of antithrombotic medications not only leads to a higher incidence of bleeding events but may also result in an increased number of CV events. However, previous studies have rarely compared bleeding risks between patients with multiple CV events and patients with general CV disease. A post hoc analysis of the results of the ISAR-REACT5 study, which primarily included patients from Germany and Italy, revealed that AMI patients with an MI history did not have an increased incidence of bleeding events during the follow-up period compared with patients with new-onset AMI.^17^ Other studies have indicated that patients at high ischemic risk, as defined using ESC/EACTS guidelines (which considers specific clinical and procedural characteristics), did not exhibit an increased incidence of clinically relevant bleeding events.^9^ By contrast, in our study, patients in the multiple-event group who were categorized as having high ischemic risk exhibited a higher rate of bleeding events. This discrepancy emphasizes the potential variability in outcomes related to bleeding risks among different racial groups. This may be significant, because many studies have indicated that patients of Asian descent treated with antiplatelet or anticoagulant medications have a higher risk of bleeding than patients of European descent.^18^ Further analysis of the timing of major bleeding events in our patients revealed that the events primarily occurred within the first 3 months following the index ACS event. This observation suggests that the bleeding events may result from the use of stronger antithrombotic medications during the acute phase of ACS. Antiplatelet or anticoagulant medications are typically prescribed for treating a recurrent CV event in the acute phase. Thus, multiple CV events might lead the treatment team to consider intensifying or prolonging the use of antiplatelet therapy. Previous studies have also indicated that patients with recurrent ACS events are more likely to have received antithrombotic agents before the ACS event.^12^

Over the 5-year follow-up period, patients in the multiple-event group exhibited a higher rate of CV death. Patients in the multiple-event group also exhibited a higher rate of non-CV death. This may be due to the higher number of comorbidities in these patients. Recent registry data from Finland indicates that an increased number of recurrent CV events not only increases the probability of new CV events but also elevates the risk of non–cardiac-related death.^19^

This study has several limitations. First, it was a retrospective cohort study that was conducted using data from a health insurance database, which lacks specific laboratory data (such as creatinine, HbA1c, and BNP levels), data on patients undergoing cardiac catheterization (e.g., presence of triple vessel CAD and final TIMI flow), and post-myocardial infarction echocardiographic data on left ventricular ejection fraction (LVEF). These factors are potentially relevant to the prognosis of ACS patients. ^20-22^ Second, the study only matched the multiple-event and single-event groups by age and sex, without further matching by other baseline characteristics, which may affect outcome assessment.

## Conclusion

Our study demonstrated that patients in the multiple-event group not only experienced increased rates of CV and non-CV mortality but also had a high incidence of both major and minor bleeding events.

## Data Availability

The data used in this study were obtained from the Taiwan National Health Insurance Research Database (NHIRD), which is managed by the National Health Research Institutes (NHRI). Access to the NHIRD is restricted and requires permission from the NHRI. Researchers who meet the criteria for access to confidential data can apply for access at the NHRI?s website (http://nhird.nhri.org.tw).

## Acknowledgments

This research was conducted with support from AstraZeneca Taiwan limited. The authors would like to acknowledge the Laboratory Animal Center at TMU for their technical support in this experiment and the assistance in revising the English manuscript.

